# Linking abdominal imaging traits to electronic health record phenotypes

**DOI:** 10.1101/2020.09.08.20190330

**Authors:** Matthew T. MacLean, Qasim Jehangir, Marijana Vujkovic, Yi-An Ko, Harold Litt, Arijitt Borthakur, Hersh Sagraiya, Mark Rosen, David A. Mankoff, Mitchell D. Schnall, Haochang Shou, Julio Chirinos, Scott M. Damrauer, Drew A. Torigian, Rotonya Carr, Daniel J. Rader, Walter R. Witschey

**Author notes:** Address correspondence to: Walter R. Witschey, Department of Radiology, Perelman Center for Advanced Medicine, 3400 Civic Center Boulevard, Philadelphia, PA 19104.

## Abstract

Quantitative traits obtained from computed tomography (CT) scans performed in routine clinical practice have the potential to enhance translational research and genomic discovery when linked to electronic health record (EHR) and genomic data. For example, both liver fat and abdominal adipose mass are highly relevant to human disease; non-alcoholic fatty liver disease(NAFLD) is present in 30% of the US adult population, is strongly associated with obesity, and can progress to hepatic inflammation, cirrhosis, and hepatocellular carcinoma. We built a fully automated image curation and organ labeling technique using deep learning to identify liver, spleen, subcutaneous and visceral fat compartments in the abdomen and extract 12 quantitative imaging traits from 161,748 CT scans in 19,624 patients enrolled in the Penn Medicine Biobank (PMBB). The average liver fat, as defined by a difference in attenuation between spleen and liver, was −6.4 ± 9.1 Hounsfield units (HU). In 135 patients who had undergone both liver biopsy and imaging, receiver operating characteristic (ROC) analysis revealed an area under the curve(AUC) of 0.81 for hepatic steatosis. The mean fat volume within the abdominal compartment for subcutaneous fat was 4.9 ± 3.1 L and for visceral fat was 2.9 ± 2.1 L. We performed integrative analyses of liver fat with the phenome extracted from the EHR and found highly significant associations with chronic liver disease/cirrhosis, chronic non-alcoholic liver disease, diabetes mellitus, obesity, hypertension, renal failure, alcoholism, hepatitis C, use of therapeutic adrenal cortical steroids, respiratory failure and pancytopenia. Liver fat was significantly associated with two of the most robust genetic variants associated with NAFLD, namely rs738409 in *PNPLA3* and rs58542926 in *TM6SF2*. Finally, we performed multivariate principle component analysis (PCA) to show the importance of each of the quantitative imaging traits to NAFLD and their interrelationships with the phenome. This work demonstrates the power of automated image quantitative trait analyses applied to routine clinical imaging studies to fuel translational scientific discovery.

## Introduction

Advanced imaging is a critical component of health care, is utilized extensively in modern medicine, and is a major component of the electronic health record (EHR). Medical centers collect enormous quantities of imaging data that could be extremely valuable for translational science, but quantitative traits are not systematically extracted for use by either researchers or clinicians. Automated measurement of quantitative traits from imaging obtained during clinical care that are then integrated with EHR and genomic data obtained via biobanking protocols at scale will fuel new biological discovery, inform disease etiology and pathophysiology, and guide the next generation of precision medicine therapies *(1)*.

Both liver fat and abdominal adipose mass are examples of quantitative traits that are highly relevant to human health and disease and can be quantitated from medical images such as CT scans *(2–6)*. Non-alcoholic fatty liver disease (NAFLD) affects approximately 30% of the US adult population and can progress to hepatic inflammation, cirrhosis, and hepatocellular carcinoma *(7, 8)*. Excess liver fat is the predominant histologic and radiologic feature that identifies NAFLD patients and is associated clinically with obesity, cardiovascular disease and diabetes *(9, 10)*. The automated extraction of liver fat and adipose mass from clinical imaging studies at scale would be of great interest for integration with other aspects of the phenome via the EHR, as well as potentially with genomic and biomarker data, to advance translational science and precision medicine.

To address the challenges of conventional analysis of large numbers of images, machine learning can be brought to bear to provide precise image analysis using automation*(11, 12)*. Deep learning has rapidly increased in popularity, becoming a dominant method in computer vision *(13)*, yet most studies have explored only a few imaging traits and do not integrate this information with the EHR to show the extent to which these traits are associated with the disease phenome *(14–17)*. An additional challenge is that patients often undergo multiple imaging scans for which machine learning has not been trained, limiting its application at scale. Our overall objective was to develop an automated approach that addresses these limitations and shows the association between multiple imaging traits and the disease phenome.

We built a fully automated image curation and organ labeling technique using deep learning applied to CT scans to identify liver, spleen, subcutaneous and visceral fat compartments in the abdomen and quantify 12 imaging phenotypes from those regions. After rigorously validating the deep learning methods, we applied it to 161,748 CT scans from 19,624 patients enrolled in the Penn Medicine Biobank (PMBB), a centralized resource of annotated blood and tissue samples linked with clinical EHR and genetic data. We performed integrative analyses of the imaging traits with other phenotypic data extracted from the EHR using principle component analysis (PCA). This work demonstrates the power of automated image quantitative trait analyses to fuel translational science by leveraging imaging studies performed in clinical care and linked to an academic biobank.

## Results

### Radiologic studies in participants in the Penn Medicine Biobank (PMBB)

In the University of Pennsylvania Health System between January 1988 and April 2019, more than 18 million radiology studies were performed, of which ∼2.7 million were CT scans (**Fig. 1A**). Approximately 50% of the CT studies (n = 1,331,012) were abdominal or chest CTs (**Fig. 1B**). At the time of this study, the PMBB had recruited ∼50,000 participants. We found that imaging in PMBB participants is extremely common: from 1988 to 2019, 170,209 CT, 96,129 magnetic resonance imaging (MRI), and 89,923 ultrasound studies were performed on patients who had at some point been enrolled in the PMBB (**Fig. 1C**). After chest x-ray, the most widely utilized advanced imaging studies performed were CT studies of the abdomen and chest, of which 64,473 were suitable for analyses (based on CPT code). Overall, 19,624 unique PMBB participants had undergone abdominal or chest CT scanning, with a total of 161,748 scans performed, providing a large sample for carrying out this study.

**Figure 1:**
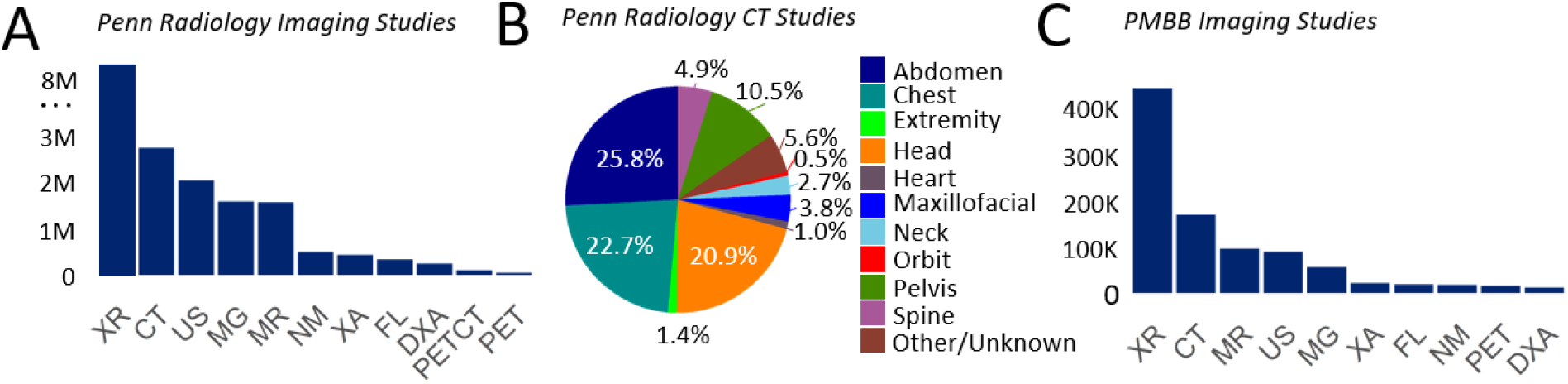
Survey of advanced imaging data in the Penn Medicine health system. **A**, Total number of stored imaging studies organized by modality, with dates ranging from 1988 to 2019. **B**, Fraction of CT scans performed by region. Abdominal and chest CT scans together comprise the two largest fractions of CT studies. **C**, Total number of imaging studies in the Penn Medicine Biobank by modality, with dates ranging from 1988 to 2019. XR = x-ray, CT = computed tomography, US = ultrasound, MR = magnetic resonance, MG = mammography, XA = x-ray angiography, FL = fluoroscopy, DXA = dual-energy x-ray absorptiometry, NM = nuclear medicine, PETCT = positron emission tomographty and computed tomography, PET = positron emission tomography.

### Automated extraction of quantitative traits from abdominal and chest CT images

Using abdominal and chest CT scans, we set about to develop an automated approach for quantifying 12 image-derived phenotypes (IDPs): liver HU (attenuation) mean, liver HU deviation, liver fat, liver volume, spleen HU mean, spleen HU deviation, spleen volume, subcutaneous fat volume, subcutaneous fat area, visceral fat volume, visceral fat area, and visceral-subcutaneous fat ratio (see Supplementary Table 1 for list of IDPs). As shown in **Fig. 2**, the fully automated technique filters images and removes non-axial orientations and high noise data such as thin sections (slice thickness < 2 mm) as detailed in methods. It identifies non-contrast or contrast enhanced CT scans (CNN_1_), identifies images that show abdominal anatomy (CNN_2_), and labels pixels showing liver (CNN_3A_), spleen (CNN_3B_), subcutaneous and visceral fat anatomy (CNN_3C_). The output of CNN_3C_ provides the inner abdominal contour which yields visceral and subcutaneous fat compartments (**Fig. 2B**). The explicit form of the CNNs appear in **Supplementary Fig. 1**.

**Figure 2:**
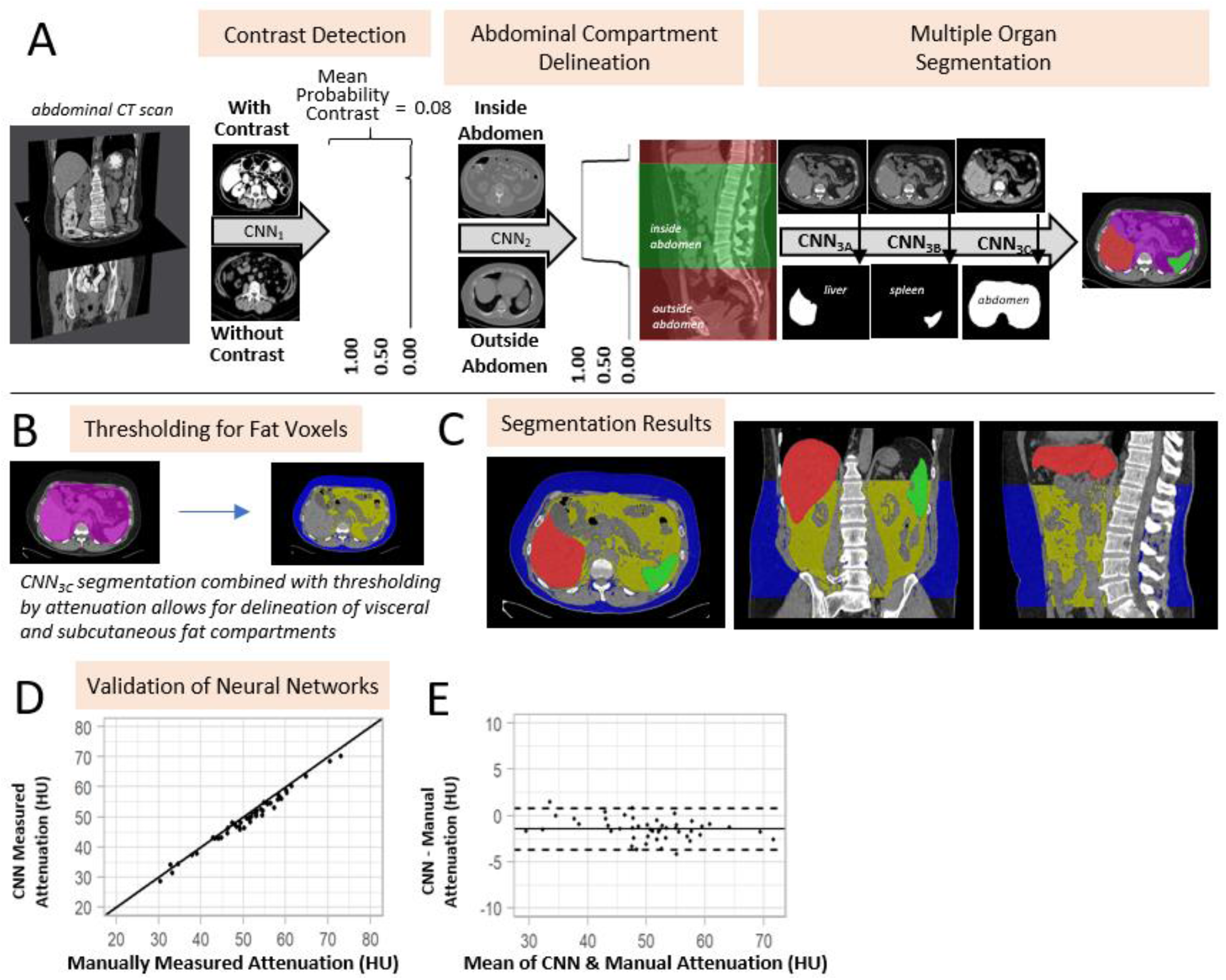
Automated extraction of quantitative imaging traits from CT scans. **A**, Patient images stored in a digital picture archiving and communication system (PACS) are queried by current procedure terminology code (a 5-digit number indicated the procedure, such as abdominal CT with contrast), anonymized and uploaded to cloud storage and computing platforms. Automated extraction of quantitative image phenotypes includes filtering to detect invalid image scans from the associated imaging metadata. A pair of convolutional neural networks detect whether the imaging scan is a contrast enhanced scan (CNN_1_) and whether an image is located inside or outside the abdomen (CNN_2_). Three sequential CNNs are used to extract liver (CNN_3A_), spleen (CNN_3B_), and abdominal (CNN_3C_) anatomy. **B**, By thresholding voxels inside and outside the abdominal segmentation, the visceral and subcutaneous fat compartments are delineated. **C**, Semantic labeling of pixels showing the liver (red), spleen (green), subcutaneous (blue) and visceral (yellow) fat structures. **D**, Agreement between expert radiologist and automatic labeling of liver volume and **E**, Bland-Altman showing mean bias. Correlation and Bland-Altman for spleen, subcutaneous and visceral fat are shown in **Supplementary Fig. 2**. BMI = body mass index, CNN = convolutional neural network, CT = computed tomography

We validated the software to measure the performance of each component (**Supplementary Table 2**). The performance of CNN_1_ to identify contrast enhanced CT scans was evaluated using 400 randomly selected scans, composed of 200 with IV contrast and 200 without. Of the 400 scans tested, 399 were classified correctly. The performance of CNN_2_ to identify the abdominal borders was assessed by randomly selecting another 100 CTs of the abdomen and pelvis and comparing the CNN and manually derived results. For the superior border, the automated method was within approximately one slice from the selected slice(1.01±1.11) and for the inferior border within one slice from the selected one (0.70±0.64). Performance of CNN_3_ in labeling liver, spleen, visceral and subcutaneous fat was found by comparing with an expert radiologic observer (D.T.) as detailed in methods. There was excellent agreement and minimal bias between all expert and automatically derived metrics as shown in **Supplementary Table 2**.

After validation, we automatically extracted 12 IDPs from abdominal and chest CT exams in the PMBB. As shown in **Fig. 3**, 161,748 CT scans were processed by our algorithm and data was extracted for 86,311 scans, corresponding to 63,160 studies, representing 19,301 patients who remained for IDP quantitation. The clinical characteristics of these patients are shown in **Table 1**. As shown in **Fig. 2A**, patient scans were retrieved from Penn Medicine picture archiving and communication server (PACS), anonymized, and image data was synchronized with a cloud computing platform for processing (Amazon Web Services). Quantitative imaging traits were extracted and mean values for each of the 12 quantitative IDPs are shown in **Table 2** and distributions of traits are shown in **Supplementary Fig. 4**.

**Table 1.** Population characteristics for cohort

| Characteristic | Full Cohort (n=19,301<sup>†</sup>) |  |
| --- | --- | --- |
| *Demographics* | *mean* | *std. dev.* |
| Age (years) | 58.1 | 15.0 |
| Sex | *n* | *%* |
| Male | 9,972 | 51.7 |
| Female | 9,312 | 48.3 |
| Ancestry |  |  |
| European | 12,461 | 64.6 |
| African | 4,609 | 23.9 |
| Asian | 307 | 1.6 |
| Other/Unknown | 1,924 | 10.0 |
| *Clinical Metrics* | *mean* | *std. dev.* |
| Body mass index, kg/m<sup>2</sup> | 28.8 | 6.9 |
| Systolic Blood Pressure, mmHg | 126.6 | 12.0 |
| Diastolic Blood Pressure, mmHg | 74.6 | 7.3 |
| *Diagnoses* | *N* | *%* |
| Hypertension |  |  |
| Yes | 10,247 | 63.7 |
| No | 5,849 | 36.3 |
| Diabetes |  |  |
| Yes | 5,176 | 34.4 |
| No | 9,865 | 65.6 |
| Heart Failure |  |  |
| Yes | 3,859 | 24.6 |
| No | 11,821 | 75.4 |
| Ischemic Heart Disease |  |  |
| Yes | 4,844 | 29.5 |
| No | 11,557 | 70.5 |
| Cerebral Vascular Accident |  |  |
| Yes | 800 | 5.2 |
| No | 14,502 | 94.8 |
| *Lab Values* | *mean* | *std. dev.* |
| Cholesterol, mg/dL | 176.0 | 38.8 |
| HDL cholesterol, mg/dL | 51.2 | 15.8 |
| LDL cholesterol, mg/dL | 98.7 | 31.9 |
| Triglycerides, mg/dL | 123.5 | 63.2 |
| HbA1c, % | 6.4 | 2.2 |
| AST, IU/L | 25.0 | 30.9 |
| ALT, IU/L | 24.9 | 33.3 |

<sup>†</sup>19,301 patients of 19,624 total (see **Fig. 3**)

**Table 2.**
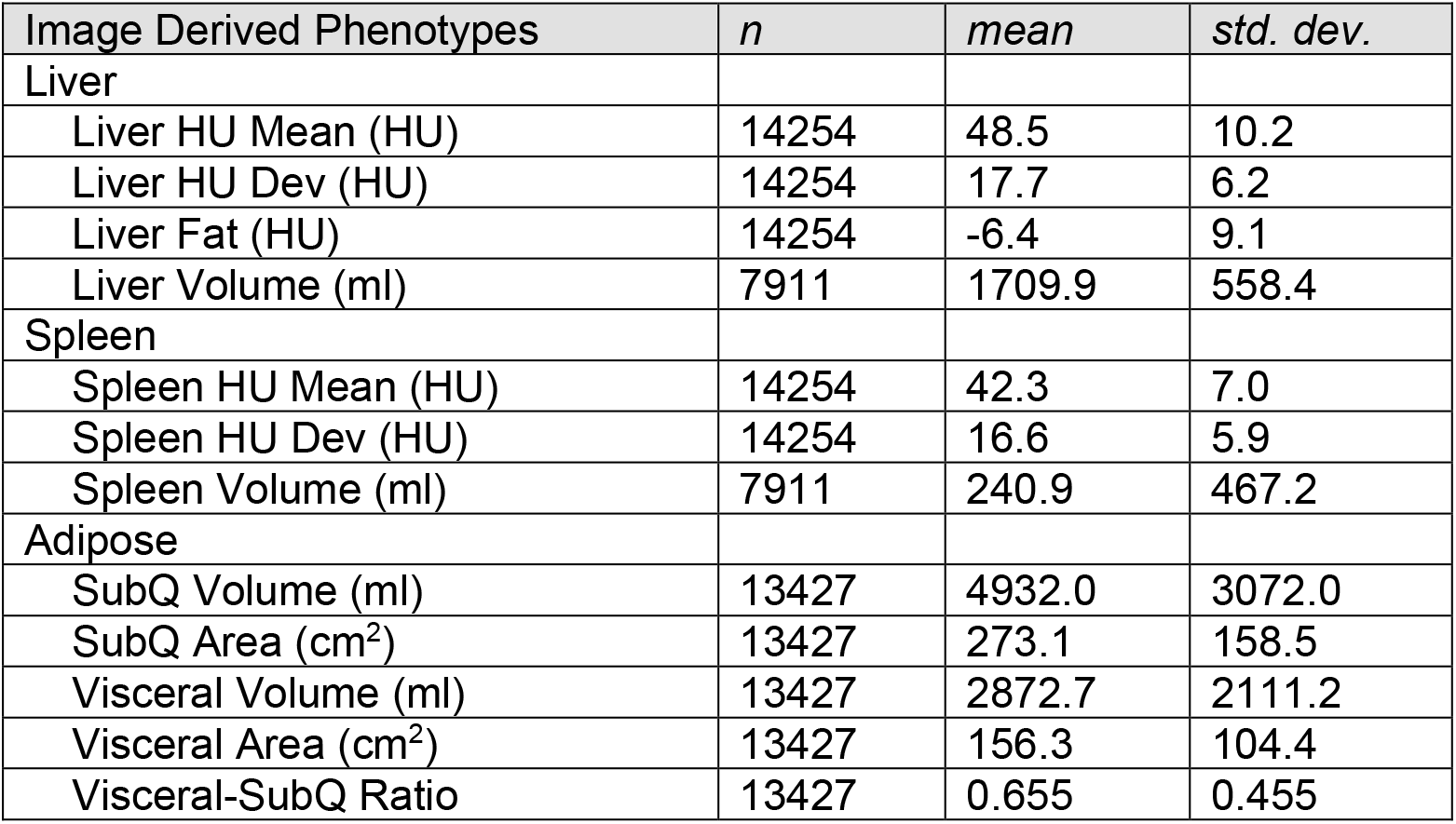
Mean values for image derived phenotypes

| Image Derived Phenotypes | *n* | *mean* | *std. dev.* |
| --- | --- | --- | --- |
| Liver |  |  |  |
| Liver HU Mean (HU) | 14254 | 48.5 | 10.2 |
| Liver HU Dev (HU) | 14254 | 17.7 | 6.2 |
| Liver Fat (HU) | 14254 | -6.4 | 9.1 |
| Liver Volume (ml) | 7911 | 1709.9 | 558.4 |
| Spleen |  |  |  |
| Spleen HU Mean (HU) | 14254 | 42.3 | 7.0 |
| Spleen HU Dev (HU) | 14254 | 16.6 | 5.9 |
| Spleen Volume (ml) | 7911 | 240.9 | 467.2 |
| Adipose |  |  |  |
| SubQ Volume (ml) | 13427 | 4932.0 | 3072.0 |
| SubQ Area (cm<sup>2</sup>) | 13427 | 273.1 | 158.5 |
| Visceral Volume (ml) | 13427 | 2872.7 | 2111.2 |
| Visceral Area (cm<sup>2</sup>) | 13427 | 156.3 | 104.4 |
| Visceral-SubQ Ratio | 13427 | 0.655 | 0.455 |

**Figure 3:**
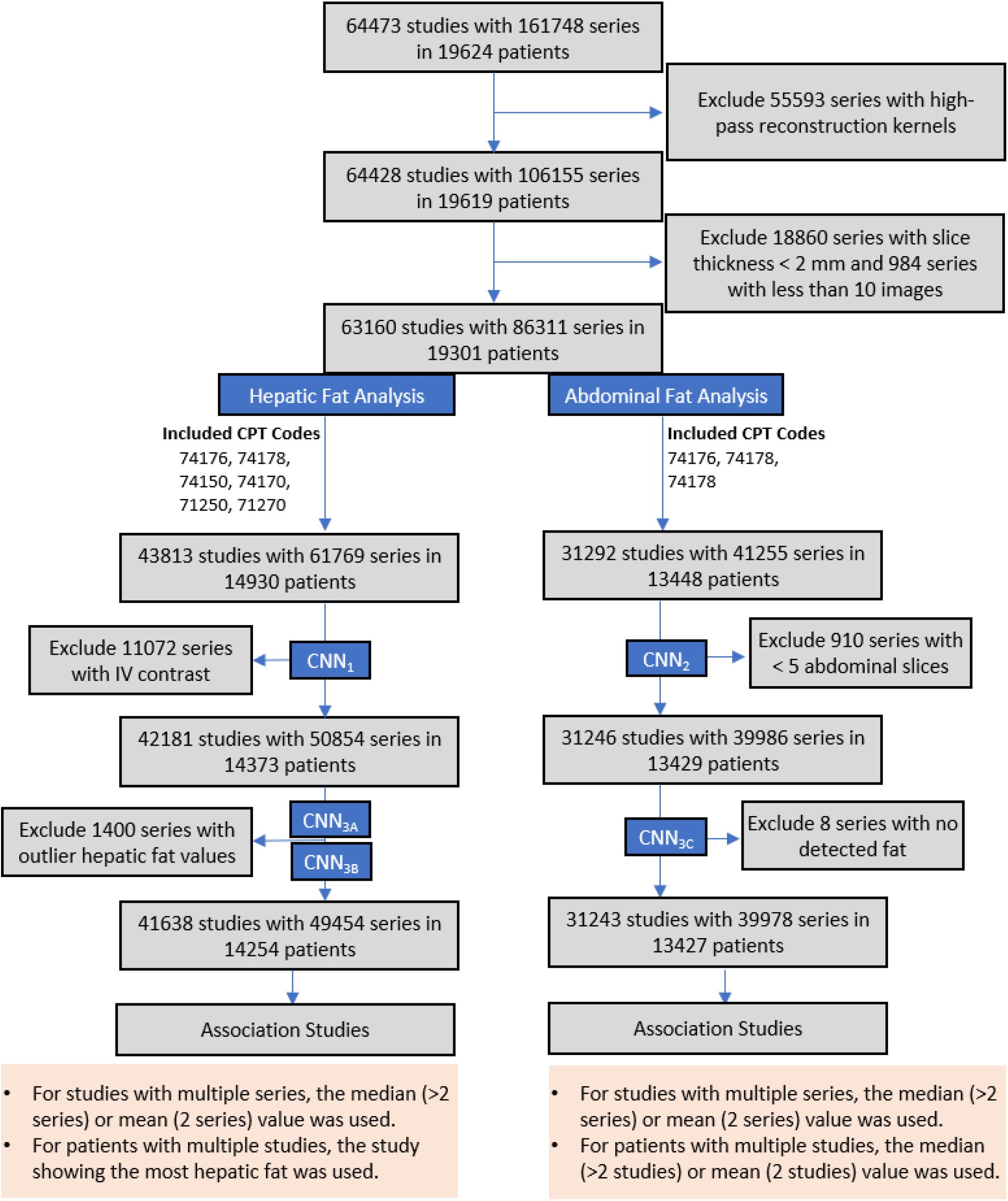
Number of patients, studies, and scans included in the study as well as the number utilized in different analyses.

### Association of liver fat with clinical traits and diagnoses

A total of 14,930 unique patients in PMBB had CT scans suitable for quantitation of liver fat and associated IDPs. Liver fat was calculated by taking the mean attenuation of the spleen and subtracting the mean attenuation of the liver; a higher value indicates more hepatic fat. After attributing a single liver fat (LF) value to each patient, LF had a mean of –6.4 ± 9.1 Hounsfield units.

144 patients with a CT scan appropriate for quantitation also had liver biopsy with a pathology report in the EHR. 94 were negative for steatosis and 50 were positive for steatosis as detailed in methods, and a receiver operator characteristic (ROC) analysis of the liver fat measurement resulted in an area under the curve (AUC) of 0.81 (**Fig. 4A**). Liver fat that provided a balance between sensitivity and specificity was −6 HU with a sensitivity of 0.66, and a specificity of 0.79. When using this threshold of −6 HU to dichotomize patients into steatosis or no steatosis, 6052 (42.5%) patients in our cohort were found to be steatotic. We then compared laboratory values between the steatotic patients (LF ≥ –6) and non-steatotic patients (LF < −6 HU) as shown in **Fig. 4B**. Steatosis was significantly associated with elevated AST (p = 3.5e-18), ALT (p = 3.4e-26), and alkaline phosphatase (p = 1.2e-9). HbA1c (p = 5.6e-4) and triglycerides (p = 1.2e-9) were significantly increased and HDL-C (p = 2.9e-14) and LDL-C (p = 1.3e-3) were decreased in patients with steatosis.

**Figure 4:**
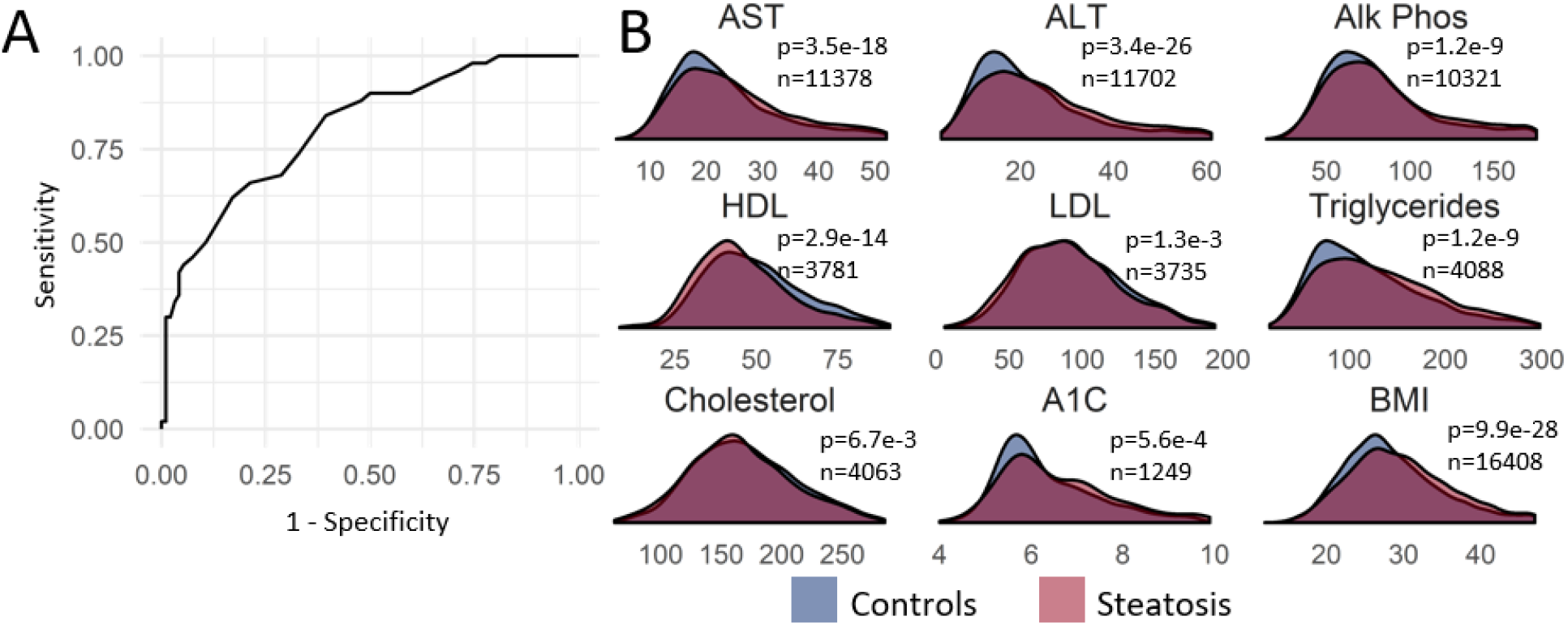
Relationship between circulating blood biomarkers, body mass, histology and hepatic fat as determined by liver CT attenuation. **A**, Receiver-operating-characteristic (ROC) for detection of liver fat on all CT from histological examinations in the PMBB. **B**, circulating blood biomarker distributions dichotomized by CT-derived liver fat (HU difference between spleen and liver = –6). AST = aspartate aminotransferase, ALT = alanine aminotransferase, Alk Phos = alkaline phosphatase, HDL = high density lipoprotein, LDL = low density lipoprotein, A1C = glycated hemoglobin, BMI = body mass index.

As expected, patients with steatosis had an elevated body mass index (BMI) (p = 9.9e-28). Because NAFLD is strongly associated with obesity, we examined the correlations between liver fat and adipose volumes. There was a significant correlation between liver fat and visceral adipose volume (r = 0.19, p< 2.2e-16) and subcutaneous fat volume (r = 0.064, p = 2.9e-9). The visceral-subcutaneous fat ratio was significantly correlated with liver fat (r = 0.15, p< 2.2e-16).

The quantitative trait of liver fat was used to perform a phenome-wide association study (PheWas) against the diagnostic codes in the EHR, in order to ask in an unbiased manner which diagnoses were disproportionately associated with increased liver fat (**Fig. 5**). There were a total of 281 diagnoses that were significantly associated with increased liver fat. Some of the strongest associations were with chronic liver disease/cirrhosis (p = 5.7e-93), chronic nonalcoholic liver disease (p = 1.9e-87), diabetes mellitus (p = 3.5e-84), obesity (p = 7.8e-42), hypertension (p = 2.4e-44), renal failure (p = 7.7e-49), alcoholism (p = 9.0e-22), hepatitis C (p = 1.1e-14), use of therapeutic adrenal corticosteroids (p = 2.1e-13), respiratory failure (p = 9.4e-35) and pancytopenia (2.3e-19). These very strong associations help to validate the liver fat quantitation and show interesting and less well-established associations of liver fat with clinical conditions.

### Association of hepatic fat with genetic variants known to be associated with NAFLD

Two well-established coding variants associated with NAFLD are rs738409 in the gene *PNPLA3* (encoding the missense variant I148M) and rs58542926 in the gene TM6SF2 (encoding the missense variant E167K) *(18)*. A total of 5,268 PMBB participants with liver fat quantitation were genotyped for these variants. We found highly significant associations with both rs738409 (MAF 0.21, p = 1.2e-11, beta = 1.49, 95% CI: 1.06–1.91) and with rs58542926 (MAF 0.06, p = 4.2e-8, beta = 2.10, 95% CI: 1.33–2.81). These significant genetic associations further validate our liver fat quantitation and suggest that with a larger sample size of genotyped and imaged individuals, this approach could be valuable in helping to identify additional genomic loci associated with NAFLD.

### Multi-feature analysis shows relatedness of image-derived phenotypes

Based on the top associations between imaging traits and phecodes, we further explored these relationships using principal component analysis (PCA). **Fig. 6A** is a PCA biplot that shows how the phecodes (black dots) are projected in the first two principal components (PCs) space by their associations with imaging traits. The arrows indicate the relative contributions of each imaging trait towards such groupings on these components. Of note, subcutaneous and visceral fat related traits all point in a similar direction, with spleen HU deviation and contribute heavily to the first PC. The phecode for sleep disorders has strong negative loadings on PC1 but almost zero loading on PC2, supporting the concept that sleep disorders such as sleep apnea are positively associated with increased adiposity. The chronic liver disease and cirrhosis phecode loads heavily on PC2, indicating a strong positive association mostly explained by imaging traits regarding liver fat trait maps. Spleen volume also maps strongly in this direction, suggesting an association with splenomegaly, which may result secondary to liver pathology. Liver fat and liver HU mean points in opposite directions because liver attenuation is inversely related to liver fat. **Fig. 6B** shows the PCA results by projecting the disease domains instead of the phecodes onto the first two PCs space of imaging traits. Consistently, digestive and hematopoietic pathologies are located where liver fat and spleen volume are most relevant, while endocrine, neurologic, and respiratory disease systems are projected to share similar patterns of association with imaging traits, mostly contributed by visceral and subcutaneous fat related traits.

**Figure 6:**
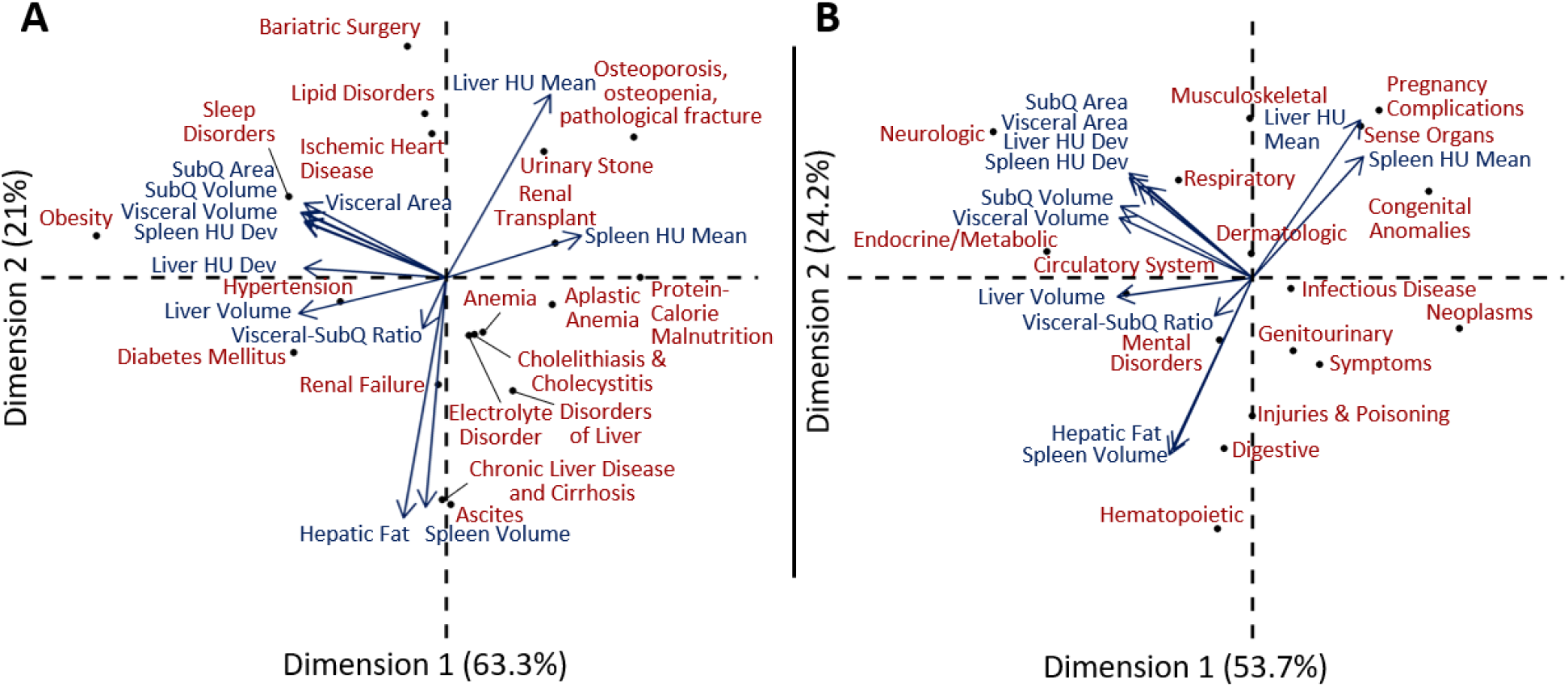
Principal component analysis representation of (**A**) phecodes and (**B**) disease systems for image derived phenotypes. Position of phecodes (**A**) and disease systems (**B**) in terms of the first two principal components is shown with points while the mapping of the original IDPs is shown with arrows.

## Discussion

Using deep learning, we extracted liver, spleen, and abdominal fat quantitative imaging traits from 19,624 patients enrolled in the Penn Medicine Biobank who had undergone clinical CT scanning. Our method was validated against liver biopsy data in a subset of patients. Using these quantitative trait imaging data and integrating them with EHR phenotype data, we found 2,495 significant associations between these imaging traits and the EHR disease phenome. Hepatic fat was very strongly associated with expected diagnoses such as obesity, diabetes mellitus, hypertension, chronic nonalcoholic liver disease, alcoholism, and cirrhosis, as well as with less obvious diagnoses such as chronic kidney disease, respiratory failure, and pancytopenia. Associations of hepatic fat with visceral adipose mass, transaminases, HbA1c, triglycerides, and genetic variants known to be associated with NAFLD provided additional confidence in the data. Multivariate analysis of imaging traits showed the extent to which each imaging trait is connected to the disease phenome; hepatic fat and spleen volume showed the strongest association with chronic liver disease. This is a proof of concept study performed at scale and these data provide a basis to pursue new or unexpected connections between liver and abdominal imaging traits and progression of disease. While several studies have employed deep learning to extract imaging traits *(11, 12, 19)*, there has been little research to integrate these traits with the EHR or an academic biobank resource at scale. In several recent studies, hepatic fat was quantified using deep learning or other algorithms *(14, 15, 20, 21)*, but was limited to liver traits or did not investigate the association between imaging traits and other types of information such as the disease phenome, genotyping data and histopathology.

Clinical CT data is heterogenous and uses different scanners, manufacturers and reconstruction algorithms. It required an autonomous and fault-tolerant approach to accurately evaluate multiple imaging traits from the diverse set of 161,748 scans. Since hepatic fat quantitation relies on scans without intravenous (IV) contrast and study metadata is often unreliable, it was necessary to separate non-contrast scans using deep learning for contrast classification. Variabilities in patient position and body habitus required a separate network to classify images that show the abdomen for accurate quantification of abdominal fat. Multiple deep learning networks were used to extract the 12 imaging traits. The approach was autonomous and can process uncurated clinical CT data. It can automatically report imaging traits in patients and show their traits with respect to patients across the biobank. The difference in spleen and liver attenuation or spleen volume showed higher association with chronic liver disease than liver attenuation alone and is explained by variations in body habitus as well as scanner type, which affect absolute liver attenuation.

Hepatic steatosis is prevalent and estimated to be present in approximately one-third of US adults; we found 42% of our patients (6,052) had steatosis, which was consistent with an older and less healthy group of patients in a tertiary health care center compared with the general population *(22)*. The value at which the spleen-liver attenuation should be dichotomized to represent steatotic and non-steatotic patients varies widely in the literature. Studies using biopsy data for validation have recommended cutoffs ranging from –3.2 to +9 *(23–25)* compared to our finding of −6 derived using ROC analysis with biopsy data. Using this cutpoint to dichotomize patients into steatosis or not, we found that steatosis was significantly associated with elevated AST, ALT, and alkaline phosphatase, consistent with findings in previous studies *(26, 27)*. Steatosis was also positively associated with HbA1c and triglycerides and negatively with HDL-C, also consistent with previous findings *(28)*.

There are several limitations to this study. Prospective studies of hepatic fat traits appear to show better accuracy against pathology *(24, 29)*. Given the allowance of up to two years, the degree of steatosis may have also fluctuated and the difference in the measurements could reflect this physiologic change. Furthermore, most of the reports had a qualitative indicator of steatotic involvement. The percent involvement that is reflected by terms such as “mild” or “moderate” steatosis is not standardized and could reflect values lower than the 30% cutoff value. The labeled liver may include vasculature, lesions, or border pixels which are traditionally excluded when measuring attenuation. Additionally, images with artifacts or organs with tumors, cysts, or pronounced vasculature could skew attenuation values. Using CT to measure liver fat has also only been shown sensitive in patients with moderate to severe steatosis indicating that we are missing a portion of the population with mild disease *(25)*. This study is also limited as phenotypes were curated from EHR billing codes and is often incomplete.

In conclusion, this study presents a new automated method for the quantification of abdominal and liver fat from clinical CT scans and shows how autonomous image trait quantification in the context of academic biobanks can facilitate genomic, biomarker, and translational research.

## Materials and Methods

### Penn Medicine Biobank

The Penn Medicine Biobank (PMBB) is a resource for integrating quantitative traits derived from clinical imaging studies with genetics, blood biomarker and other electronic health record data at Penn Medicine, a multi-hospital health system headquartered in Philadelphia, PA. PMBB is a detailed long-term, prospective, epidemiological study of over 50,000 volunteers containing approximately 27,485 diagnostic codes (ICD9 and ICD10). It is ethnically diverse and African-Americans represent approximately 25% of the patients. At present, the BioBank does not integrate data from advanced imaging, yet more than 18 million imaging studies were performed by Penn Medicine since 1998 and 3 million radiology studies in 2018 and 2019 alone making this an underutilized BioBank resource. All patients provided informed consent to participate in the PMBB and to utilization of electronic health record and image data.

### Study Design

Within our biobank cohort of 52,441 patients, we queried the EHR image-server based on Current Procedural Terminology (CPT) codes for all non-contrast chest (CPT 71250, 71270), non-contrast abdomen (CPT 74150, 74170), and all abdomen/pelvis (CPT 74176, 74177, 74178) CT scans. Chest studies were included as these routinely include one-third to one-half of the liver, and an even larger fraction of the spleen, which is sufficient for evaluation of liver fat. Based on this query, we identified 64,473 studies with a total of 161,748 axial image scans representing 19,624 patients. Scans using high-pass reconstruction kernels, such as Siemens I50F, were excluded as these are intended for edge detection tasks and create high-degrees of image noise. Additionally, scans with slice thickness less than 2 mm were excluded as these are known to introduce noise. Finally, image stacks with less than 10 images were excluded as these likely represent incomplete datasets. Liver fat analysis was conducted on all non-contrast chest, abdomen, and abdomen/pelvis studies corresponding to CPT codes 71250 (n = 26,959), 71,270 (n = 1116), 74150 (n = 2,383), 74170 (n = 1,410), 74176 (n = 10,031), 74178 (n = 1,914). In total, 43,813 studies containing a total of 61,769 image scans representing 14,930 patients were processed for liver fat analysis. Measurement of visceral and subcutaneous fat was performed on all abdominal/pelvis studies corresponding to CPT codes 74176 (n = 10,031), 74177 (n = 19,347), and 74178 (n = 1,914). In total 31,292 studies containing a total of 41,255 image scans corresponding to 13,448 patients were processed for visceral and subcutaneous fat analysis. All training data was manually generated by a trained technician under the supervision of a board-certified abdominal radiologist using 3D Slicer software. Application of exclusion criteria as well as the number of scans processed in each part of the study is shown in **Fig. 3**.

We developed deep learning algorithms for one of the most widely used imaging modalities, computed tomography (CT), to automatically extract quantitative traits pertaining to abdominal and liver fat from > 14,000 patients. By using deep learning to automatically extract CT image traits, we performed large-scale associations with phenotyping (N>13,000) and genomic (N>5,000) data to provide automated assessment of fatty liver disease and potential underlying conditions.

### Image Analysis

#### Hardware and Image Prefiltering

All architectures were implemented in Python using the Tensorflow package in the cloud (Amazon Web Services). Training was conducted using an NVIDIA P100 graphical processing unit (GPU) and inferences used parallel processing across 8 NVIDIA K80 GPUs. The network inputs were 2D axial slices with size of 256×256 pixels and were thresholded with a window width of 150 Hounsfield units (HU) and a level of 30 HU for the contrast classification network (CNN_1_) as well as liver (CNN_3A_) and spleen (CNN_3B_) segmentation networks. A window width of 1800 HU and level of 400 HU was used for the abdominal compartment delineation network (CNN_2_) and a window width of 400 HU and a level of 50 HU was used for the abdominal contour segmentation network (CNN_3C_). See **Fig. 2** for a depiction of each network’s role in our project.

#### Model Architecture – Detection of Non-Contrast CT Scans

The first network (CNN_1_) identified intravenous (IV) contrast CT scans and removed them from the liver fat analysis pipeline. Images were shown to convolutional layers which flattened into fully connected layers modeled after the VGG-16 classification network [11] (**Supplementary Fig. 1A**). This architecture showed excellent performance for labeling of images in the ImageNet competition. The network outputs a probability between 0 and 1 indicating the likelihood that the slice contains IV contrast. Scans were considered to have contrast if the average per-slice probability was greater than 0.5. This network was trained on 800 scans, 400 with IV contrast, and 400 without. Additionally, half these scans were of the abdomen/pelvis and half were thoracic. Of the abdomen/pelvis scans, within the IV contrast group 158 contained additional PO contrast and within the no-IV contrast group 115 contained PO contrast. 320 patients (50,654 slices) were randomly placed in the training group, and 80 patients (12,867 slices) in a validation group. Training was conducted over 10 epochs with batch-size of 32. A separate testing set was randomly selected from the Penn Medicine Biobank and contained 50 abdomen/pelvis studies and 50 thoracic studies. To evaluate performance of VGG classification network an additional 400 scans were randomly selected from PMBB data with 200 being thoracic and 200 of the abdomen/pelvis. Of the abdomen/pelvis scans, 75 of the IV contrast scans also had PO contrast and of those without IV contrast, 44 contained PO contrast. See **Supplementary Fig. 2** for a summary of the scan types used in testing CNN_1_.

#### Model Architecture – Identification of Images Showing Abdominal Anatomy

The second network (CNN_2_) labeled 2D slices as belonging to the abdominal cavity. This network standardized the quantification of subcutaneous and visceral fat across patients by identifying defined anatomical endpoints for the abdominal cavity. The same network architecture that was used in IV contrast classification (CNN_1_) was used for this task of labeling abdominal cavity images. The network was trained to output a probability between 0 and 1 indicating the likelihood that the slice is within the boundary of the abdomen. The superior border of the abdomen is defined as the first slice in which the lungs are no longer visible. The inferior border of the abdomen is defined as the first slice that the bottom of the L5 vertebra is visible. These borders were manually labeled on 468 training scans and the network was trained to differentiate between two classes – the abdominal slices (located within the borders) and the not-abdominal slices (located outside the borders). Of these scans, 375 were used for model training and 93 for validation. Within the training group, there were 13536 abdominal slices and 21769 not-abdominal slices. Within the validation group, there were 3374 abdominal slices and 5401 not-abdominal slices. Training was conducted over 30 epochs with a batch size of 32. A testing set of 100 abdomen/pelvis CT scans was selected at random from the PMBB on which the performance of the automated model was compared to manual labeling.

#### Model Architecture – Segmentation

Additional networks were trained to segment the liver, spleen, subcutaneous and visceral fat from axial 2D slices modeled after a U-Net architecture *(30)*. This model is composed of symmetric paths joined by skip connections where localized feature information from the contracting path is combined with contextual information from the expanding path. The complete architecture is shown in **Supplementary Fig. 1B**. The networks output a probability for each voxel indicating the probability that it belongs to the foreground. For liver segmentation, the network (CNN_3A_) was trained on a total of 106 scans with 81 scans (7,999 slices) randomly selected for training and 25 scans (2,436 slices) for validation. For spleen segmentation, the network (CNN_3B_) was trained on a total of 158 scans with 127 scans (12,399 slices) randomly selected for training and 31 scans (2,865 slices) for validation. For abdominal compartment segmentation, the network (CNN_3C_) was trained on a total of 50 scans with 40 scans (1627 slices) randomly selected for training and 10 scans (430 slices) for validation. Training data was selected iteratively when the model underperformed on a scan. Of note, to identify subcutaneous and visceral fat, we do not train a network to directly provide these segmentations. Instead, we first segment the abdominal compartment (see CNN_3C_ in **Fig. 2A** and **Fig. 2B**), and then locate fat voxels by thresholding for attenuation values between –190 and −30. All fat voxels within the abdominal compartment segmentation are visceral fat and all those outside the segmentation are subcutaneous fat. Training was conducted with a batch-size of 32 over 135 epochs for liver, 108 for spleen, and 106 for the abdominal compartment network.

To evaluate segmentation performance, a testing set of 20 abdomen/pelvis CT scans was randomly selected from PMBB and both manual as well as automated segmentations for liver and spleen were produced. DICE coefficients were calculated to measure agreement between manual and automatic segmentations. Additionally, 50 scans were selected at random and mean attenuation was measured in the liver and spleen by the manual placement of ROIs. Eight spherical (20 mm diameter) ROIs were placed in the liver with four in the left and four in the right lobe. Two spherical (15 mm diameter) ROIs were placed in the spleen. Care was taken to avoid placement near edges or in regions of vasculature or lesions. The mean HU was computed between ROIs for the liver and spleen and compared to that obtained from the automated approach. To evaluate performance of the abdominal compartment network, the abdominal compartment was manually contoured in a testing set of 10 randomly selected abdomen/pelvis CT scans and DICE coefficients were calculated to measurement agreement between manual and automatic segmentations. Additionally, visceral/subcutaneous fat measures were manually acquired on a single slice between L3 and L4 on 100 randomly selected abdomen/pelvis scans and then compared with automatically derived metrics.

### Image Derived Phenotypes

A total of 12 image derived phenotypes (IDP) were quantified on a subset of the total 64,473 CT studies. These IDPs describe volume or attenuation properties of organs based on the results of our image processing networks. The IDPs are defined as follows:

#### Liver fat

liver fat (LF) was quantified by subtracting the mean attenuation of all voxels contained within the liver from the mean attenuation of all voxels contained in the spleen. This IDP was computed on all non-contrast chest, abdomen, abdomen&pelvis scans (CPT 74176, 74178, 74150, 74170, 71250, 71270) for a total of 41,638 studies representing 14,254 patients.

#### Liver HU Mean

calculated by taking the mean attenuation of all voxels contained within the liver segmentations. This IDP was computed on the same subset of scans as *liver fat*.

##### Liver HU Dev

calculated by taking the standard deviation of attenuation for all voxels contained within the liver segmentations. This IDP was computed on the same subset of scans as *liver fat*.

##### Spleen HU Mean

calculated by taking the mean attenuation of all voxels contained within the spleen segmentations. This IDP was computed on the same subset of scans as *liver fat*.

##### Spleen HU Dev

calculated by taking the standard deviation of attenuation for all voxels contained within the spleen segmentations. This IDP was computed on the same subset of scans as *liver fat*.

##### Liver Volume

calculated by computing the volume in milliliters for all of the liver segmentations. This IDP was computed on all non-contrast abdomen, abdomen&pelvis CT scans (CPT 74176, 74178, 74150, 74170) for a total of 14,572 studies representing 7,911 patients.

##### Spleen Volume

calculated by computing the volume in milliliters for the spleen segmentations. This IDP was computed on the same subset of scans as *liver metric volume*.

##### SubQ (Subcutaneous) Volume

calculated by computing the volume in milliliters for the subcutaneous fat region. This IDP was computed on all abdomen&pelvis CT scans (CPT 74176, 74177, 74178) for a total of 31,243 studies representing 13,427 patients.

##### Visceral Volume

calculated by computing the volume in milliliters for the visceral fat region. This IDP was computed on the same subset of scans as *subq volume*.

##### SubQ Area

calculated by computing the mean per-slice area of subcutaneous fat in cm^2^ for all scans in the abdominal compartment. This IDP was computed on the same subset of scans as *subq volume*.

##### Visceral Area

calculated by computing the mean per-slice area of visceral fat in cm^2^ for all scans in the abdominal compartment. This IDP was computed on the same subset of scans as *subq volume*.

##### Visceral-SubQ Ratio

calculated by taking the ratio of *SubQ Area and Visceral Area*. This IDP was computed on the same subset of scans as *subq volume*.

For a given IDP, each patient often has multiple measurements as each patient may have multiple studies and each study multiple scans. In order to conduct association studies, it is necessary to assign a single IDP value to each patient. For the *liver fat* and *liver HU mean* IDPs, when a study had multiple scans, the scan with a value closest to the median value was taken for studies with 3 or more scans and closest to the mean for those with 2 scans. For patients with multiple studies, the maximum value of *liver fat* and the minimum value for *liver HU mean* was taken across all studies. While liver attenuation in a patient can vary with time, we wanted to capture values for patients that represented the highest degree of fatty infiltration. For this reason, we took the maximum *liver fat* value and the minimum *liver HU mean* across all studies. For all other IDPs, when handling both a study with multiple scans or a patient with multiple studies, the median (greater than 3) or mean (exactly 2) values were utilized.

#### Biopsy Specimen Validation

To validate the ability of our automated method to detect liver fat, liver biopsy reports were analyzed to identify patients who had pathology proven steatosis. A total of 1,124 pathology reports were reviewed for patients who also had CT scans. Biopsy reports that indicated a history of or plans for a transplant were excluded as this would complicate pairing with the appropriate CT study. Reports indicating only mild steatosis were also excluded as CT is considered diagnostic only for steatosis of greater than 30% involvement. If the report listed a percent steatosis, reports were categorized based on this percentage with values of greater than 30% being positive and those less than 5% being negative for steatosis. Intermediate values in the range of 5% to 30% were excluded. In the absence of a definitive measurement, cases with qualitative terms such as “mild” or “minimal” were excluded while those indicating “moderate”, “severe”, or “marked” steatosis were considered positive for greater than 30% involvement. Next, the scans were each matched to the nearest pathology report. If no reports existed for a given patient or if there was no scan-report pair less than 2 years apart, those patients’ scans were excluded. One scan-report pair with the smallest difference in dates was then kept for each remaining patient. To evaluate performance of our liver fat metric in assessing steatosis, receiver operating characteristic (ROC) analysis was performed. Area under the ROC curve was measured and a balanced cutoff value was selected by choosing the Liver fat threshold that resulted in the coordinate nearest to the upper left corner.

#### Phenome-wide association study

A phenome-wide association study (PheWAS) was performed to investigate the phenotypic associations of fatty liver disease. ICD10 codes were first mapped to ICD9 codes using the 2017 general equivalency mapping (GEM). Next, ICD9 codes were aggregated into phecodes using the PheWAS R package to create 1,816 phecodes. Patients with at least 2 occurrences of a PheCode are considered cases, those with none are controls, and those with 1 are treated as missing. Phecodes with less than 100 cases were excluded. The ICD9 code 571.8 is used for patients with non-alcoholic fatty liver disease *(31, 32)*.

#### Genome-wide association study

Within the PMBB, there are 19,515 total genotype samples from three different genotype arrays: Illumina Omni Express (n = 10,867), Infinium GSA-Version 1.0 (n = 5,676), and Infinium GSA-Version 2.0 (n = 2,972). Data from each array was quality-controlled (QC) separately. QC steps included identifying sex mismatches, applying marker (< 95%) and sample (< 90%) call-rate filters, and removing duplicates. Prior to imputation, strand alignment was performed and consisted of matching SNPs to the appropriate positive strand file on 1000 Genomes reference panel, removing palindromic SNPs and removing SNPs absent from reference panel *(33, 34)*. Additionally, PLINK (version 2.0) was used to identify and flip all SNPs where the top allele was not on the positive strand *(35)*. SNPs were removed where strands could not be flipped, and the allele frequency differed > 40% from whites in 1000 Genomes *(36)*. Imputation was performed for all autosomes on Michigan Imputation Server with the Haplotype Reference Consortium selected as the reference panel *(37)*. Phasing was conducted using EAGLE *(38)* and imputation was performed using MINIMAC software *(34)*. After imputation, the genotype data was merged together. Relatedness was assessed using a graph-based algorithm after applying a Pi-hat threshold of 0.25 to account for relatives up-to first cousins to be removed. One sample with the highest degree of relatedness from a list of related samples was removed from the dataset (1616 total samples). The top 20 principal components were computed using SmartPCA on a genotype dataset that included all PMBB participants and an additional 2,504 individuals from 1000 Genomes *(36)*. These PCs were then used for to determine genetically inferred ancestry and compose ancestral groups used for population stratification into European and African American groups. Subsequent PMBB-specific PCA analysis was performed, for which the first 4 PCs were included as covariates in the regression model.

A genome-wide association study (GWAS) was conducted on the subset of 5,268 patients with both a non-contrast CT scan and genotyping data. This GWAS was conducted on all ancestries including European (n = 3,102), African (n = 1,964), and other (n = 202). Linear regression was used to test for associations between SNPs and the continuous phenotype of liver fat, controlling for sex, age, age^2^, ancestry, and the first four principal components.

#### Statistical Analysis

- **Fig. 1D, 1E, Supplementary Fig. 2**: In all regression plots, *y = x* is shown as a solid line and the line of best fit as a dotted line. In the bland-altman plots, the solid arrow shows the mean value and the dotted lines show the range of the first standard deviation.
- **Fig. 4**: Lab values acquired greater than 90 days and BMI measurements acquired greater than 365 days from the scan-date were excluded. A Wilcoxon rank-sum test was performed to assess if lab values for steatotic vs. non-steatotic patients were significantly different.
- **Fig. 5** (PheWas): Logistic regression was performed with each phecode as the outcome and liver fat as a predictor. Regression was performed controlling for the covariates of age, sex, and race. Bonferroni multiple comparison correction was used to determine the level of significance.
- **Fig. 6** (PCA): To generate **Fig. 6**, we first grouped phecodes. Phecodes were rounded to the nearest whole number and then grouped using a logical ‘and’ operation. For example, the phecode 250 is diabetes mellitus, 250.1 is type 1 diabetes, and 250.2 is type 2 diabetes. Under our grouping system, all three of these phecodes were reduced to a single phecode 250 which was positive if any of the three phecodes were positive. Logistic regression was performed for each grouped phecode as the outcome and each IDP as a predictor while controlling for the covariates of age, sex, and race. Grouped phecodes with less than 100 cases were excluded. We recorded the z-scores for each of these regressions. After grouping and exclusion, there were a total of 317 phecodes remaining. Next, for each IDP, we retained the top 5 phecodes with the largest z-score (absolute value) for a total of 19 phecodes. We then performed principal component analysis (PCA) on these phecodes by traits z-score matrix as a dimension-reduction method to visualize and group the phecodes jointly based on the phecode-IDP association patterns. **Fig. 6A** shows the position of phecodes projected onto the first two principal component space by IDP, and the relatively contributions (arrows from origin) of each IDP to the two principal components. **Fig. 6B** was generated in a similar manner. However, instead of grouping phecodes by rounding their numeric values, we grouped all phecodes by disease system as categorized by the PheWAS R package. The figure shows the position of disease domains projected onto the first two principal component space by IDP, and their relatively contributions (arrows from origin).

**Figure 5:**
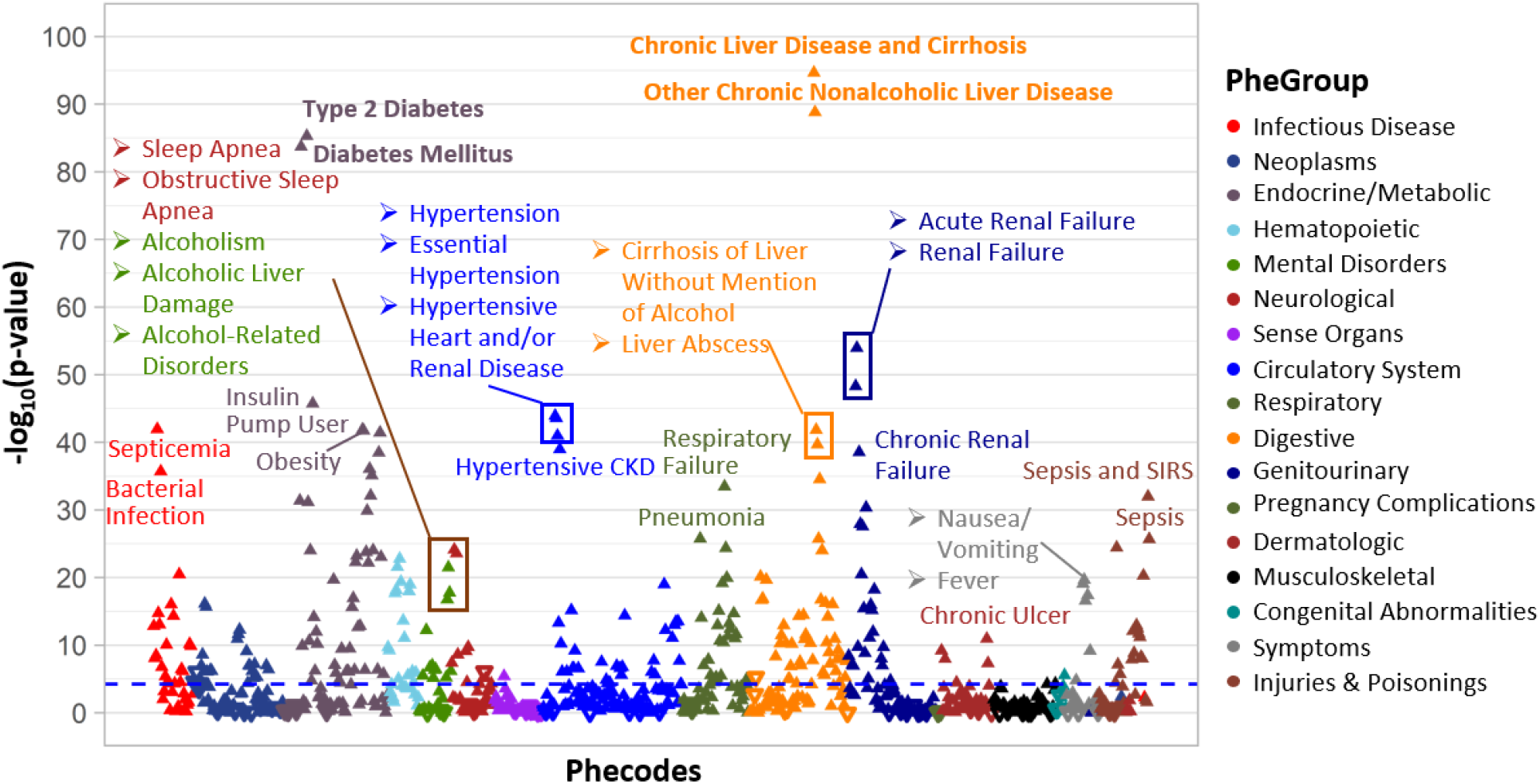
Phenome-Wide Association Study of liver fat. Blue line indicates level of significance with Bonferroni multiple-comparison correction. Upward-facing triangles indicate a positive association with increased liver fat and downward-facing triangles indicate a negative association.

When analyzing associations between laboratory values and liver fat, patients were first dichotomized into steatotic or non-steatotic using a LF threshold of −6. This value of −6 was the balanced cutoff value obtained from the ROC analysis using biopsy data. A LF value greater than −6 implies that the spleen has an attenuation that is either within 6 Hounsfield Units less than that of the liver or greater than that of the liver. In cases where there were multiple lab values for a given patient, the value acquired nearest to the diagnostic CT scan was selected.

## Data Availability

Data is available upon reasonable request.

## Acknowledgements

This work was supported by the Sarnoff Cardiovascular Research Foundation (MM), NIH NCATS UL1TR001878, NIH/NHLBI R01 HL137984, R01 AA026302–02, P30 DK0503060 (RC), and the Penn Center for Precision Medicine.

## SUPPLEMENTARY DATA

**Supplementary Figure 1:**
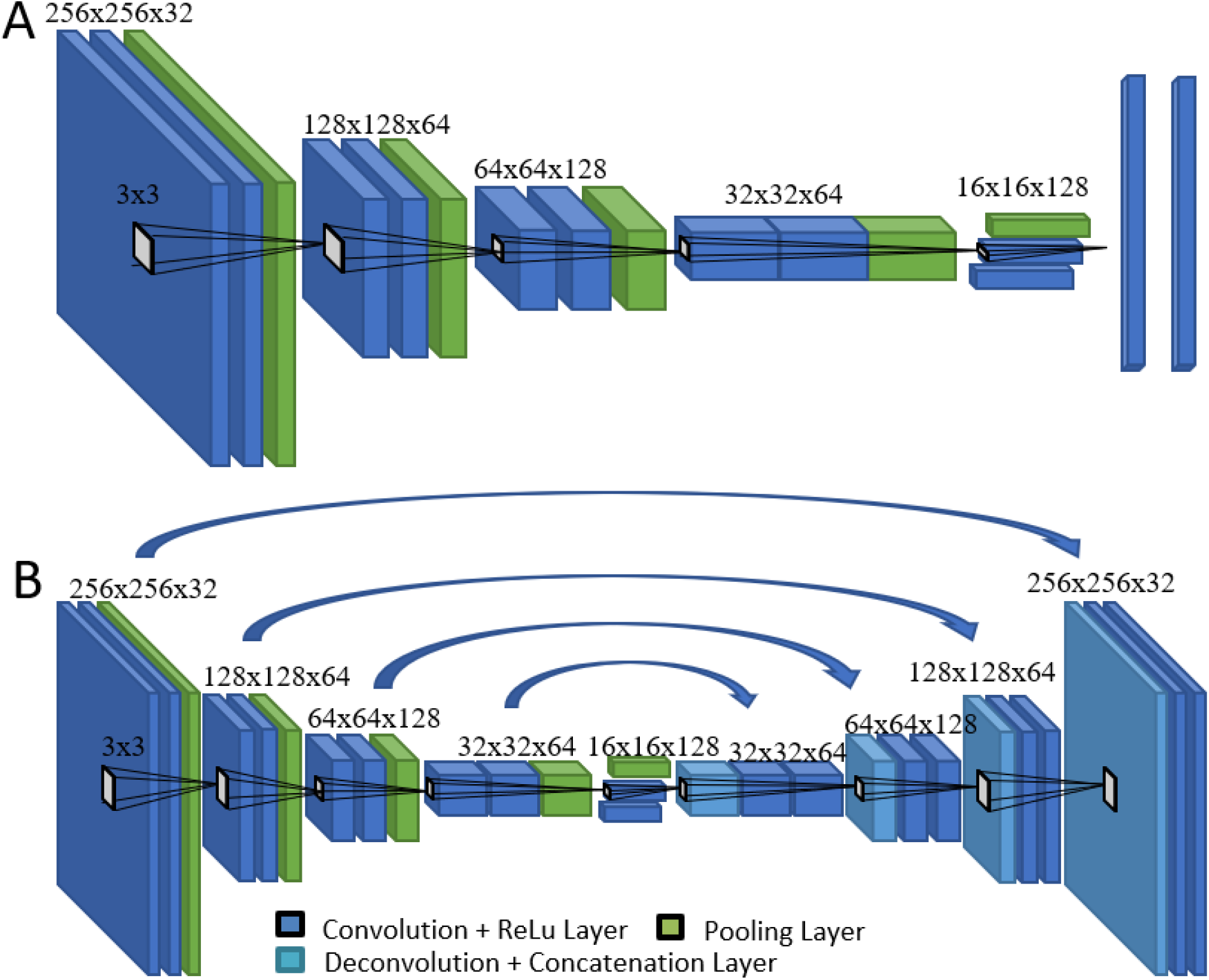
Convolutional neural networks used for labeling of **A**, contrast or non-contrast CT scans (CNN_1_) and abdominal borders (CNN_2_) and **B**, pixel-level labeling (segmentation) of liver (CNN_3A_), spleen (CNN_3B_), and subcutaneous and visceral fat (CNN_3C_). The neural network in A shows 5 stages of CNN paired with a single dense, hidden layer and with a 2 –category output layer. The CNN in B is a U-Net model network with 27 total layers in contracting and expanding paths with skip connections between layers of the same size. The final output layer is a probability map for each pixel belonging to the quantitative imaging trait.

**Supplementary Figure 2:**
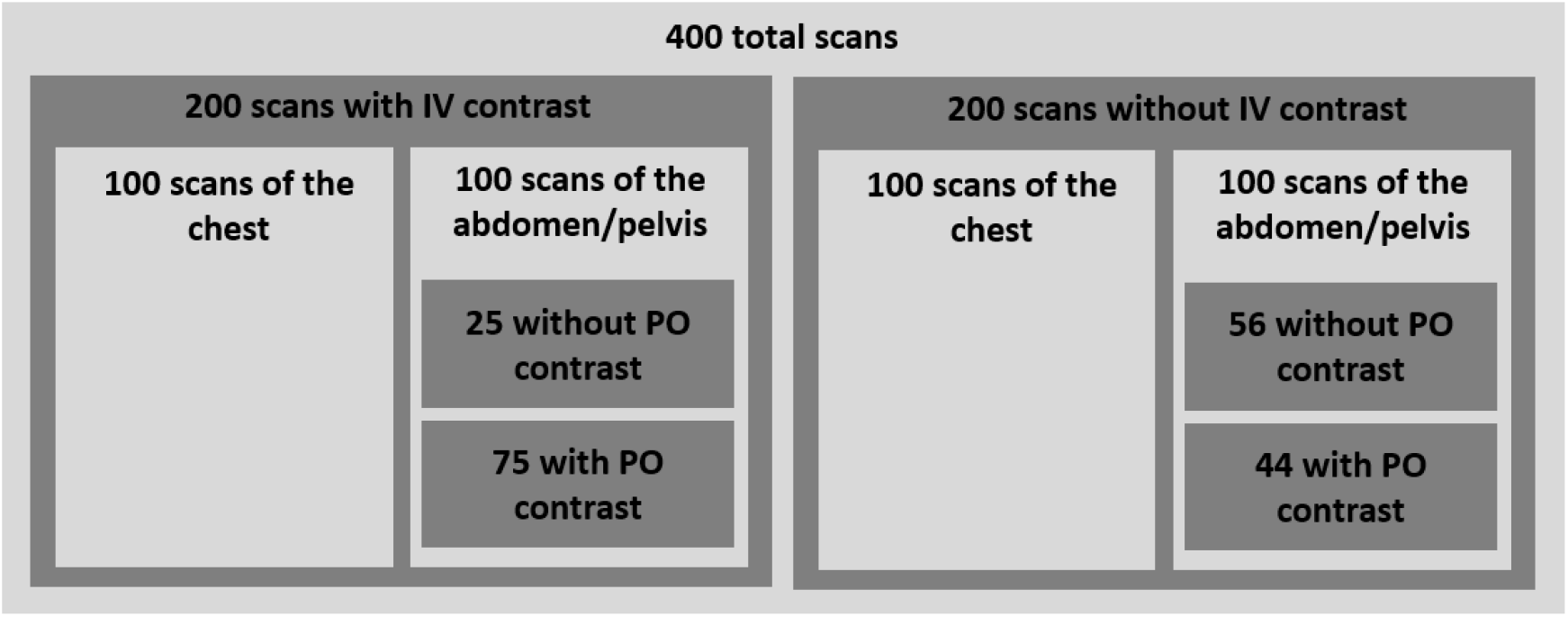
Diagram showing the number of scans within each category that were used in testing the contrast-detection network (CNN_1_).

**Supplementary Figure 3:**
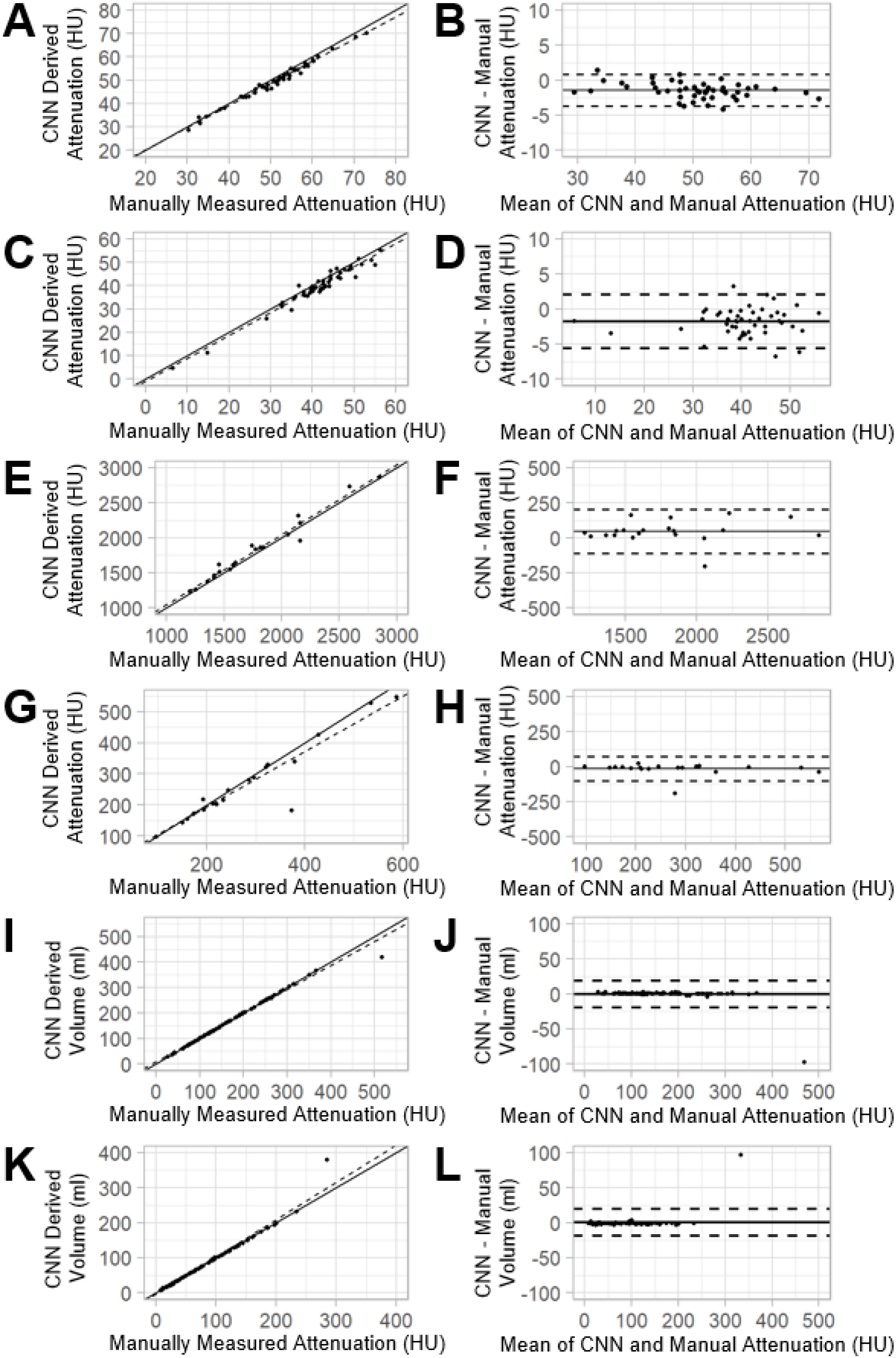
Correlation and Bland-Altman plots comparing CNN to manually derived metrics for liver Hounsfield units (A,B), spleen Hounsfield units (C,D), liver volume (E,F), spleen volume (G,H), subcutaneous fat volume (I,J), and visceral fat volume (K,L)

**Supplementary Figure 4:**
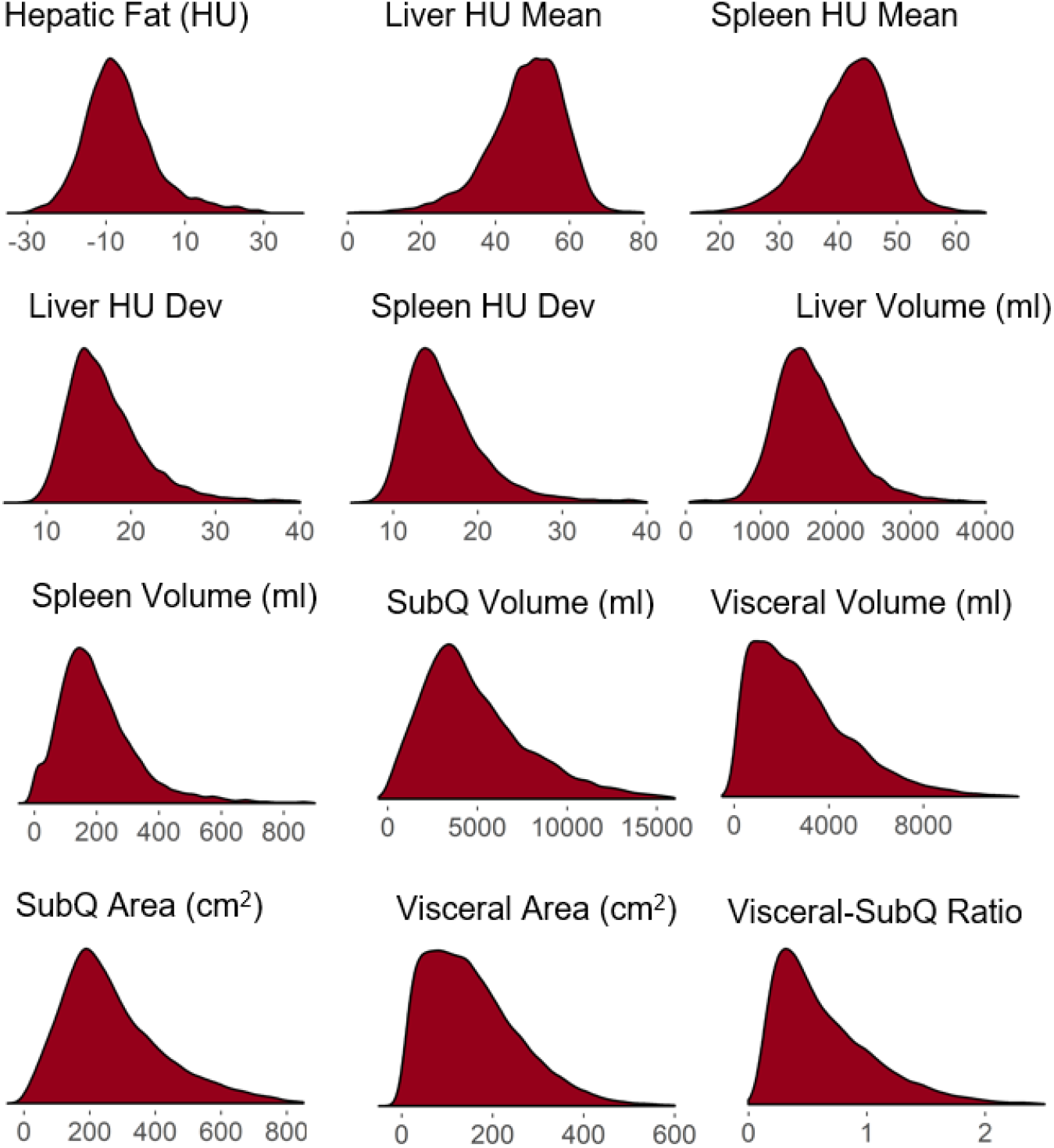
Distribution of image-derived phenotypes for biobank population.

**Supplementary Table 1.** Image Derived Phenotypes that were quantified on PMBB scans.

| Image Derived Phenotypes |
| --- |
| Liver HU Mean (HU) |
| Liver HU Dev (HU) |
| Liver Fat (HU) |
| Liver Volume (ml) |
| Spleen HU Mean (HU) |
| Spleen HU Dev (HU) |
| Spleen Volume (ml) |
| SubQ Volume (ml) |
| SubQ Area (cm<sup>2</sup>) |
| Visceral Volume (ml) |
| Visceral Area (cm<sup>2</sup>) |
| Visceral-SubQ Ratio |

**Supplementary Table 2.** Accuracy of Deep Learning Methods for Quantitative CT Traits

|  |  |  |  |  |  |
| --- | --- | --- | --- | --- | --- |
| **CNN<sub>1</sub> (Contrast Detection)** | *n* |  |  |  |  |
| Sensitivity | 400 | 100% |  |  |  |
| Specificity | 400 | 99.5% |  |  |  |
| **CNN<sub>2</sub> (Abd. Image Detection, N=100)** | *n* | *Mean* | *std. dev.* | *range* | *p* |
| Inferior position |  |  |  |  |  |
| †Bias (number of slices) | 100 | 0.70 | 0.64 | [-2, 3] |  |
| Superior position |  |  |  |  |  |
| †Bias (number of slices) | 100 | 1.01±1.11 | 1.11 | [-7, 4] |  |
| **CNN<sub>3</sub> Segmentation** | *n* |  |  |  |  |
| ‡Dice Score Coefficients |  | *mean* | *std. dev.* |  |  |
| Liver (CNN<sub>3A</sub>) | 20 | 0.95 | 0.02 |  |  |
| Spleen (CNN<sub>3B</sub>) | 20 | 0.92 | 0.07 |  |  |
| Abdominal compartment (CNN<sub>3C</sub>) | 10 | 0.98 | 0.01 |  |  |
| Subcutaneous fat (CNN<sub>3C</sub>) | 10 | 1.00 | 0.00 |  |  |
| Visceral fat (CNN<sub>3C</sub>) | 10 | 0.99 | 0.01 |  |  |
| Volume (mL): CNN vs. Manual |  | *mean bias* | *std. dev.* | *ICC* | *p* |
| Liver (CNN<sub>3A</sub>, whole volume) | 20 | 46.2 | 80.1 | 0.98 | 1.4e-15 |
| Spleen (CNN<sub>3B</sub>, whole volume) | 20 | 15.3 | 43.6 | 0.94 | 9.2e-11 |
| Subcutaneous fat (CNN<sub>3C</sub>, single slice) | 100 | 0.3 | 9.8 | 0.99 | <2e-16 |
| Visceral fat (CNN<sub>3C</sub>, single slice) | 100 | 0.3 | 9.8 | 0.99 | <2e-16 |
| Attenuation (HU): CNN vs. Manual |  | *mean bias* | *std. dev.* | *ICC* | *p* |
| Liver (CNN<sub>3A</sub>) | 50 | 1.5 | 1.3 | 0.98 | <2e-16 |
| Spleen (CNN<sub>3B</sub>) | 50 | 1.8 | 2.0 | 0.96 | <2e-16 |

†Bias indicates mean number of slices between expert label of most inferior (superior) slice position and CNN-derived estimate of that slice.
‡Dice score coefficients (DSC)= $\frac{2|X \cap Y|}{|X| + |Y|}$ , where X is the expert label and Y is the CNN label.
\*ICC = intraclass correlation coefficient

## References

1. C. J. Herold et al., Imaging in the Age of Precision Medicine: Summary of the Proceedings of the 10th Biannual Symposium of the International Society for Strategic Studies in Radiology. Radiology 279, 226–238 (2016).

2. J. L. Rehm et al., Proton density fat-fraction is an accurate biomarker of hepatic steatosis in adolescent girls and young women. Eur Radiol 25, 2921–2930 (2015).

3. B. K. Kang et al., Hepatic fat quantification: a prospective comparison of magnetic resonance spectroscopy and analysis methods for chemical-shift gradient echo magnetic resonance imaging with histologic assessment as the reference standard. Invest Radiol 47, 368–375 (2012).

4. A. Tang et al., Nonalcoholic fatty liver disease: MR imaging of liver proton density fat fraction to assess hepatic steatosis. Radiology 267, 422–431 (2013).

5. V. Bhat et al., Quantification of Liver Fat with mDIXON Magnetic Resonance Imaging, Comparison with the Computed Tomography and the Biopsy. J Clin Diagn Res 11, TC06–TC10 (2017).

6. P. J. Pickhardt et al., Specificity of unenhanced CT for non-invasive diagnosis of hepatic steatosis: implications for the investigation of the natural history of incidental steatosis. Eur Radiol 22, 1075–1082 (2012).

7. A. R. Araujo, N. Rosso, G. Bedogni, C. Tiribelli, S. Bellentani, Global epidemiology of non-alcoholic fatty liver disease/non-alcoholic steatohepatitis: What we need in the future. Liver Int 38 Suppl 1, 47–51 (2018).

8. Z. M. Younossi et al., Global epidemiology of nonalcoholic fatty liver disease-Metaanalytic assessment of prevalence, incidence, and outcomes. Hepatology 64, 73–84 (2016).

9. Q. M. Anstee, G. Targher, C. P. Day, Progression of NAFLD to diabetes mellitus, cardiovascular disease or cirrhosis. Nat Rev Gastroenterol Hepatol 10, 330–344 (2013).

10. A. Lonardo, S. Ballestri, G. Marchesini, P. Angulo, P. Loria, Nonalcoholic fatty liver disease: a precursor of the metabolic syndrome. Dig Liver Dis 47, 181–190 (2015).

11. A. Esteva et al., Dermatologist-level classification of skin cancer with deep neural networks. Nature 542, 115–118 (2017).

12. V. Gulshan et al., Development and Validation of a Deep Learning Algorithm for Detection of Diabetic Retinopathy in Retinal Fundus Photographs. Jama 316, 2402–2410 (2016).

13. M. T. Hagan, H. B. Demuth, M. H. Beale, O. De Jesús, Neural network design. (Martin Hagan, 2014).

14. P. M. Graffy, V. Sandfort, R. M. Summers, P. J. Pickhardt, Automated Liver Fat Quantification at Nonenhanced Abdominal CT for Population-based Steatosis Assessment. Radiology 293, 334–342 (2019).

15. P. F. Christ et al., in International Conference on Medical Image Computing and Computer-Assisted Intervention. (Springer, 2016), pp. 415–423.

16. Z. Guo et al., Liver Fat Content Measurement with Quantitative CT Validated against MRI Proton Density Fat Fraction: A Prospective Study of 400 Healthy Volunteers. Radiology 294, 89–97 (2020).

17. G. Chlebus et al., Reducing inter-observer variability and interaction time of MR liver volumetry by combining automatic CNN-based liver segmentation and manual corrections. PLoS One 14, e0217228 (2019).

18. B. Kahali, B. Halligan E. K. Speliotes, Insights from Genome-Wide Association Analyses of Nonalcoholic Fatty Liver Disease. Semin Liver Dis 35, 375–391 (2015).

19. B. Ehteshami Bejnordi et al., Diagnostic Assessment of Deep Learning Algorithms for Detection of Lymph Node Metastases in Women With Breast Cancer. Jama 318, 2199–2210 (2017).

20. D. E. T. Brosch, H. Schulz, S. Renisch, H. Nickisch, V. Groza, paper presented at the Medical Imaging with Deep Learning, The Netherlands: Amsterdam, 2018.

21. M. G. Linguraru, J. K. Sandberg, Z. Li, F. Shah, R. M. Summers, Automated segmentation and quantification of liver and spleen from CT images using normalized probabilistic atlases and enhancement estimation. Med Phys 37, 771–783 (2010).

22. J. D. Browning et al., Prevalence of hepatic steatosis in an urban population in the United States: impact of ethnicity. Hepatology 40, 1387–1395 (2004).

23. Y. S. Park et al., Biopsy-proven nonsteatotic liver in adults: estimation of reference range for difference in attenuation between the liver and the spleen at nonenhanced CT. Radiology 258, 760–766 (2011).

24. S. H. Park et al., Macrovesicular hepatic steatosis in living liver donors: use of CT for quantitative and qualitative assessment. Radiology 239, 105–112 (2006).

25. S. S. Lee et al., Non-invasive assessment of hepatic steatosis: prospective comparison of the accuracy of imaging examinations. J Hepatol 52, 579–585 (2010).

26. D. Sanyal et al., Profile of liver enzymes in non-alcoholic fatty liver disease in patients with impaired glucose tolerance and newly detected untreated type 2 diabetes. Indian J Endocrinol Metab 19, 597–601 (2015).

27. M. W. Pantsari, S. A. Harrison, Nonalcoholic fatty liver disease presenting with an isolated elevated alkaline phosphatase. J Clin Gastroenterol 40, 633–635 (2006).

28. M. Tomizawa et al., Triglyceride is strongly associated with nonalcoholic fatty liver disease among markers of hyperlipidemia and diabetes. Biomed Rep 2, 633–636 (2014).

29. S. W. Lee et al., Unenhanced CT for assessment of macrovesicular hepatic steatosis in living liver donors: comparison of visual grading with liver attenuation index. Radiology 244, 479–485 (2007).

30. O. Ronneberger, P. Fischer, T. Brox, U-Net: Convolutional Networks for Biomedical Image Segmentation. arXiv: 1505.04597, (2015).

31. N. Chalasani et al., The diagnosis and management of nonalcoholic fatty liver disease: Practice guidance from the American Association for the Study of Liver Diseases. Hepatology 67, 328–357 (2018).

32. R. M. Carr, A. Oranu, V. Khungar, Nonalcoholic Fatty Liver Disease: Pathophysiology and Management. Gastroenterol Clin North Am 45, 639–652 (2016).

33. G. P. Consortium, A global reference for human genetic variation. Nature 526, 68–74 (2015).

34. B. Howie, C. Fuchsberger, M. Stephens, J. Marchini, G. R. Abecasis, Fast and accurate genotype imputation in genome-wide association studies through pre-phasing. Nat Genet 44, 955–959 (2012).

35. S. Purcell et al., PLINK: a tool set for whole-genome association and population-based linkage analyses. Am J Hum Genet 81, 559–575 (2007).

36. N. Patterson, A. L. Price, D. Reich, Population structure and eigenanalysis. PLoS genetics 2, (2006).

37. S. Das et al., Next-generation genotype imputation service and methods. Nature genetics 48, 1284–1287 (2016).

38. P. R. Loh, P. F. Palamara, A. L. Price, Fast and accurate long-range phasing in a UK Biobank cohort. Nat Genet 48, 811–816 (2016).

